# Exploring the Potential of Antidepressants in Treating Coronary Artery Disease

**DOI:** 10.1101/2025.08.11.25333473

**Authors:** Yaojiang Wang, Zhidie Jin, Yuerong Jiang, Keji Chen

**Author notes:** **Correspondence:** Yuerong Jiang. These authors share first authorship (Yaojiang Wang, Zhidie Jin).

## Abstract

**Background:** Coronary artery disease (CAD) is a leading cause of death, and depression exacerbates CAD. Antidepressants may offer therapeutic potential for CAD.

**Methods:** We employed Mendelian Randomization (MR), summary-based MR (SMR), colocalization, replication analysis, and single-cell RNA annotations to assess causal relationships between antidepressant targets and CAD. Safety profiles were evaluated using the Food and Drug Administration’s (FDA) Adverse Event Reporting System (FAERS).

**Results:** Fifteen proteins demonstrated significant associations with CAD. GM2A (odds ratio [OR]: 0.975, *P* = 4 × 10⁻³), PYGL, BCHE, and several others were found to reduce the risk of CAD, while PDE4A (OR: 1.183, *P* < 1 × 10⁻³) and others were associated with an increased risk. GM2A passed sensitivity analyses and exhibited strong colocalization (posterior probability of colocalization [PPH.4] > 0.8). Elevated expression of GM2A consistently showed an inverse association with CAD risk across six tissue types, with cell-type-specific patterns observed in endothelial cells and macrophages. In SMR, FOLH1 was identified as a replicable protective factor for CAD. The FAERS recorded 52,952 adverse events (AEs) related to the selected antidepressant, affecting 6,391 patients. The predominant AEs included drug withdrawal syndrome, dizziness, paresthesia, and nausea. Significant safety signals were identified for dysphoria (reporting odds ratio [ROR] 708.12) and affect lability (ROR 362.05). Additionally, unexpected events such as insomnia, anxiety, fatigue, irritability, headache, and agitation were noted.

**Conclusions:** Our findings suggest that antidepressants may have a therapeutic role in the treatment of CAD, with GM2A identified as a promising target for therapy. While certain antidepressants can influence CAD risk, further validation is necessary to address safety concerns.

## 1 Introduction

Coronary artery disease (CAD) remains the leading global cause of mortality, driven by atherosclerotic plaque in epicardial arteries (1). Its clinical spectrum ranges from stable angina to ischemia induced by macrovascular stenoses, microvascular dysfunction, or vasospasm. Current pharmacological treatments, particularly antiplatelet and anticoagulant therapies, elevate bleeding risk, urging exploration of novel interventions targeting endothelial dysfunction and atherosclerosis prevention (2).

Recent evidence highlights the high comorbidity and complex interplay between depression and CAD (3). Depression independently increases CAD morbidity and mortality through mechanisms including poor medication adherence, platelet dysfunction, systemic inflammation, and autonomic/neuroendocrine dysregulation, accelerating atherosclerosis (4, 5). Clinical evidence suggests that treating depression may reduce CAD incidence and improve survival (6). However, some antidepressants, such as cymbalta—a serotonin-norepinephrine reuptake inhibitor (SNRI)—may carry cardiovascular risks (7). Traditional Chinese Medicine (TCM) approaches, including treatments like Xiaoyao San, demonstrate potential but require rigorous validation. The limited randomized controlled trials and ethical constraints impede definitive conclusions on antidepressant efficacy in CAD.

Given CAD’s multifactorial nature and complex antidepressant effects, robust causal inference methods are required to evaluate therapeutic potential. Mendelian randomization (MR) leverages genetic variants as instrumental variables to infer causality, mitigating confounding and reverse causation biases inherent in observational data (8). By simulating randomization at conception, MR provides lifetime effect estimates analogous to randomized controlled trials (RCTs) (9). The increasing availability of genome-wide association study (GWAS) and quantitative trait locus (QTL) data facilitates two-sample MR to assess drug-target relationships efficiently and economically. Colocalization analysis further strengthened causal inference by accounting for linkage disequilibrium and disentangling shared causal variants (10).

Proteins encoded by human genes are critical mediators of disease pathogenesis and pharmacological intervention. Drug targets with genetic links to diseases exhibit higher approval rates (11). Protein quantitative trait loci (pQTLs) enhance target discovery by more directly linking protein expression to disease phenotypes than expression QTLs (eQTLs) (12). Integrating large-scale GWAS data with pQTLs via summary-data-based Mendelian randomization (SMR) allows detection of pleiotropic protein-trait associations while differentiating pleiotropy from linkage through the Heterogeneity In Dependent Instruments (HEIDI) test (13).

Previous MR studies have assessed the relationship between general antidepressant use and CAD risk but lacked drug-specific and target-based analysis, limiting mechanistic insights (7). In this study, we address these gaps by performing a comprehensive MR investigation of antidepressant-CAD associations using multiple curated databases—ETCM, DrugBank, EpiGraphDB, deCODE, UKB-PPP—and the IEU Open GWAS Project. We apply two-sample MR, SMR, sensitivity analysis, replication analysis, Bayesian colocalization, and single-cell transcriptomic annotations to explore underlying causal pathways.

Additionally, post-marketing safety of cymbalta was assessed using the U.S. FDA Adverse Event Reporting System (FAERS), the world’s largest database of spontaneous adverse drug event reports. Employing disproportionality analysis with multiple algorithms (ROR, PRR, BCPNN, EBGM), we identified significant drug-adverse event signals.

In this study, our integrated approach identifies plasma proteins linked to CAD that are modulated by antidepressant drugs and clarifies their roles in disease pathophysiology. We also provide a detailed pharmacovigilance profile of cymbalta, elucidating potential risks. This work bridges cardiovascular and neuropsychiatric research, illustrating the promise of drug repurposing to enhance CAD management and patient outcomes. The flowchart of our study design is depicted in Fig. 1.

**Fig. 1.**
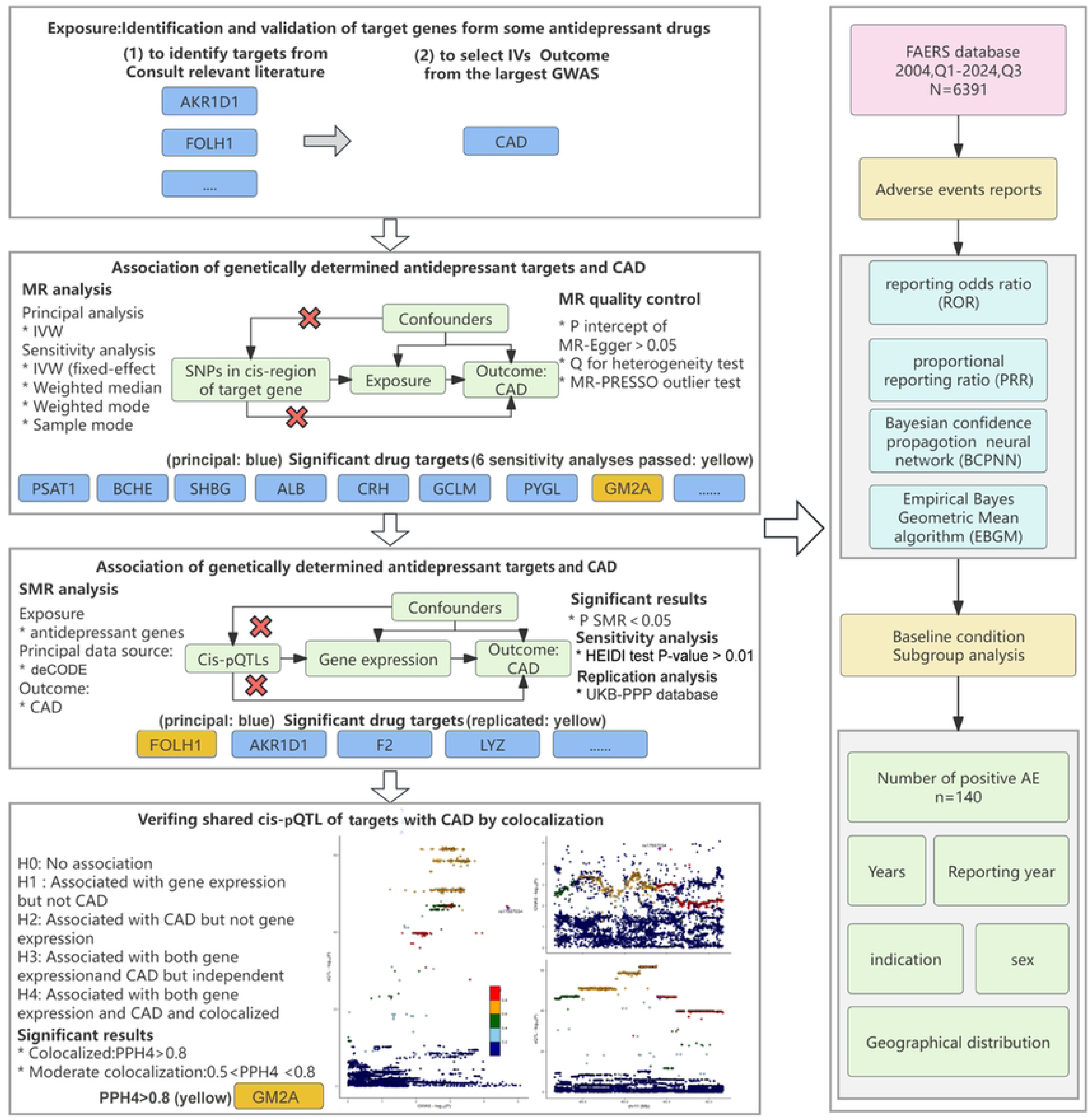
Flowchart of our study design.

## 2 Materials and methods

### 2.1 Identification of targets for antidepressants

The identification of antidepressant drug targets was conducted in four steps. First, six clinically recommended patented Chinese medicines with sufficient evidence were selected according to the *Clinical Application Guidelines for Traditional Chinese Medicine in the Treatment of Depressive Disorders* (2022): Shugan Jieyu Capsules (SJC), St. John’s Wort (*Hypericum perforatum* L.) (SJW), Morinda officinalis oligosaccharides (MOO), Shugan Granules (SGG), Xiaoyao Pills (XP), and Wuling Capsules (WC). Next, active compounds and their target genes (n = 497, accessed October 2024) were retrieved from CNKI, Wanfang databases, and the Encyclopedia of Traditional Chinese Medicine (ETCM, http://www.tcmip.cn/ETCM/). Subsequently, 28 antidepressant drugs covering major classes—including SSRIs, NaSSAs, melatonin receptor agonists, serotonin receptor modulators, NDRIs, tricyclic antidepressants (TCAs), and tetracyclic antidepressants—along with 106 pharmacologically active protein targets and encoding genes were identified through DrugBank (https://go.drugbank.com/, accessed October 2024). Finally, we compiled and deduplicated the targets from both traditional Chinese and Western medicines.

### 2.2 Gene expression datasets used for genetic instrument selection

Since pQTL directly reflect protein expression levels and the primary targets of drug action, we selected pQTL over eQTL as primary instrumental variables. We obtained pQTL data from EpiGraphDB (https://docs.epigraphdb.org/web-ui/pqtl/, accessed January–March 2024), which currently includes 1,740 plasma proteins across 576 European phenotypes. Inclusion criteria for pQTLs were (a) genome-wide significance (*P* < 5 × 10⁻⁸); (b) independent association based on LD clumping (*r*² < 0.2), using “EUR” as the LD reference panel; (c) located within 500 kb of the transcription start site of the protein-coding gene (cis-pQTL); and (d) minor allele frequency (MAF) > 0.01. Instrument strength was evaluated by the F-statistic, with F > 10 used to exclude weak instruments.

### 2.3 Outcome dataset

Genetic predictors for coronary artery disease CAD were obtained from the IEU OpenGWAS Project (trait: Coronary artery disease; dataset: ebi-a-GCST005194; https://gwas.mrcieu.ac.uk/datasets/ebi-a-GCST005194/; catalogue number: ebi-a-GCST005194; van der Harst and Verweij (14)). This GWAS meta-analysis included a total sample size of 296,525 individuals, comprising 34,541 cases and 261,984 controls of European ancestry, with summary statistics for 7,904,237 SNPs. To avoid sample overlap and minimize type I error, exposure and outcome datasets were strictly non-overlapping, ensuring robust MR analysis.

### 2.4 Two-sample MR analysis

We conducted a two-sample MR analysis to investigate causal relationships between antidepressant-associated proteins (exposures) from the EpiGraphDB database and CAD outcomes. The validity of MR causal estimates rests on three core assumptions: (1) instrumental variables (IVs) are strongly associated with the exposure; (2) IVs are independent of confounders; and (3) IVs influence the outcome solely through the exposure, with no evidence of horizontal pleiotropy.

For proteins with single pQTLs, causal estimates were derived through Wald ratio analysis. Multi-SNP instruments underwent random-effect inverse-variance weighted (IVW) regression as the primary analytical method, supplemented by fixed-effects IVW, MR-Egger, weighted median, simple mode, and weighted mode approaches. The IVW method was prioritized due to its robustness.

We quantified instrumental heterogeneity using Cochran’s Q statistic (*P* < 0.05 indicating significant heterogeneity) and assessed horizontal pleiotropy through MR-Egger intercept analysis. The MR-Pleiotropy RESidual Sum and Outlier (MR-PRESSO) method identified and corrected for outlier SNPs through its global test (significance threshold *P* < 0.05). Sensitivity analysis incorporated weighted median (effective with <50% invalid instruments) and weighted mode estimators (reliable when valid IVs constitute the modal effect cluster).

False discovery rate (FDR) correction (*q* < 0.05) controlled for type I error inflation, with significant associations (adjusted *P* < 0.05) considered suggestive evidence. Causal effects were expressed as odds ratios (OR) per standard deviation increase in protein concentration, accompanied by 95% confidence intervals. Instrument strength was validated through F-statistic calculation, excluding variants with F < 10 to mitigate weak instrument bias.

All analyses were executed in R v4.1.1 using the TwoSampleMR packages. Diagnostic visualizations included funnel plots (symmetry assessment), scatter plots (directional pleiotropy evaluation), and leave-one-out sensitivity plots.

This multi-methodological framework ensures robust causal inference while systematically addressing potential violations of MR assumptions through comprehensive sensitivity testing.

### 2.5 SMR analysis

Further SMR analysis was conducted using pQTL from the deCODE database and GWAS summary data, applying the SMR method (version 1.3.1) to estimate the causal effects of genetically predicted protein expression on CAD risk.

The analysis focused on cis-pQTLs within a 500 Kb window around each gene, primarily considering the top associated SNP. A significance level of *P* < 0.05 was set for the SMR analysis. To distinguish pleiotropic effects from linkage, the HEIDI test was applied when over three SNPs were available; a HEIDI *P*-value > 0.01 indicated the association was unlikely due to LD. Allele harmonization and all statistical tests were performed using SMR software. Python 3.12.0 was utilized to graphically represent the outcomes.

### 2.6 Colocalization analysis

LD may introduce confounding in MR studies. To address this, we applied Bayesian colocalization analysis with the *coloc* R package, assessing whether association signals for potential druggable genes and CAD-related traits share causal variants within ± 100 Mb windows. The method uses approximate Bayes factors to calculate posterior probabilities (PP) across five hypotheses: (i) H0: No association of SNPs with either trait; (ii) H1: Association of SNPs with trait 1 only; (iii) H2: Association of SNPs with trait 2 only; (iv) H3: Associations of SNPs with both traits but driven by distinct variants; and (v) H4: Associations of SNPs with both traits driven by a shared causal variant. We guide the analysis by default prior probabilities (p1 = 1×10-4 for trait 1, p2 = 1×10-4 for trait 2, and p12 = 1×10-5 for shared causality). Strong colocalization was defined as Posterior Probability of Hypothesis 4 [PP.H4] ≥ 0.8, and moderate as 0.5 < PP.H4 < 0.8. Proteins with strong, moderate, and low colocalization evidence were classified as tier 1, tier 2, and tier 3 targets, respectively. Finally, we visualized the outcomes using the *LocusCompare* R package and Python 3.12.0.

### 2.7 Replication analysis

To further validate the robustness of our SMR findings, we conducted replication analysis utilizing whole blood pQTL data from the UK Biobank Pharma Proteomics Project (UKB-PPP)—a large-scale initiative profiling the plasma proteome of 54,219 UK Biobank participants in collaboration with 13 biopharmaceutical companies.

Significant targets were identified based on principal IVW analysis. Among these, key targets were defined as those that also withstood robust sensitivity analysis (weighted median, weighted mode, simple mode), satisfied strict MR quality control thresholds (F statistic > 10, FDR < 5%, *P*_Q-stat_ > 0.05, MR-Egger intercept *p* > 0.05, MR-PRESSO *p* > 0.05), and were required to be supported by at least one positive result of SMR or colocalization analysis.

Furthermore, we further explored the tissue-specific expression and regulation of the key target genes by integrating eQTL data from the latest Genotype-Tissue Expression (GTEx) database and performing SMR analysis on key targets with available cis-eQTLs. Additionally, to delineate gene expression distributions and specificities across tissues and cell types, we leveraged single-cell transcriptomic annotations from the Human Protein Atlas (HPA), a Sweden-based program aimed at mapping human proteins across various biological contexts using diverse omics technologies.

### 2.8 Analysis of FAERS database

#### 2.8.1 Data source and data processing

To evaluate the potential risks and adverse events (AEs) associated with antidepressant therapy in CAD, we analyzed cymbalta-related AEs using data from the FAERS spanning Q1 2004 (cymbalta’s initial FDA approval) to Q3 2024. Cymbalta, a representative antidepressant, was identified in FAERS via a comprehensive search of all related brand and generic names.

Reports included those with cymbalta as the primary suspect drug. Data extraction and processing were performed in R 4.4.1. To ensure accuracy, duplicate reports were removed according to FDA guidelines: first, exact duplicates were deleted; second, among reports with the same CASEID, only the latest by FDA_DT (receipt date) was retained; third, for identical CASEID and FDA_DT, the highest PRIMARYID was kept. AEs and drug indications were coded using MedDRA 27.0 and medication data standardized using the WHO Drug Dictionary.

#### 2.8.2 Data analysis

In our study, disproportionality analysis was conducted to assess the association between cymbalta and AEs in the FAERS database, utilizing four established algorithms: Reporting Odds Ratio (ROR), Proportional Reporting Ratio (PRR), Bayesian Confidence Propagation Neural Network (BCPNN), and Multi-Item Gamma-Poisson Shrinker (MGPS). The equations and significance criteria for each algorithm are detailed in Table 1. For signal identification, an AE was considered potentially associated with cymbalta if it met the significance threshold in all methods.

**Table 1.**
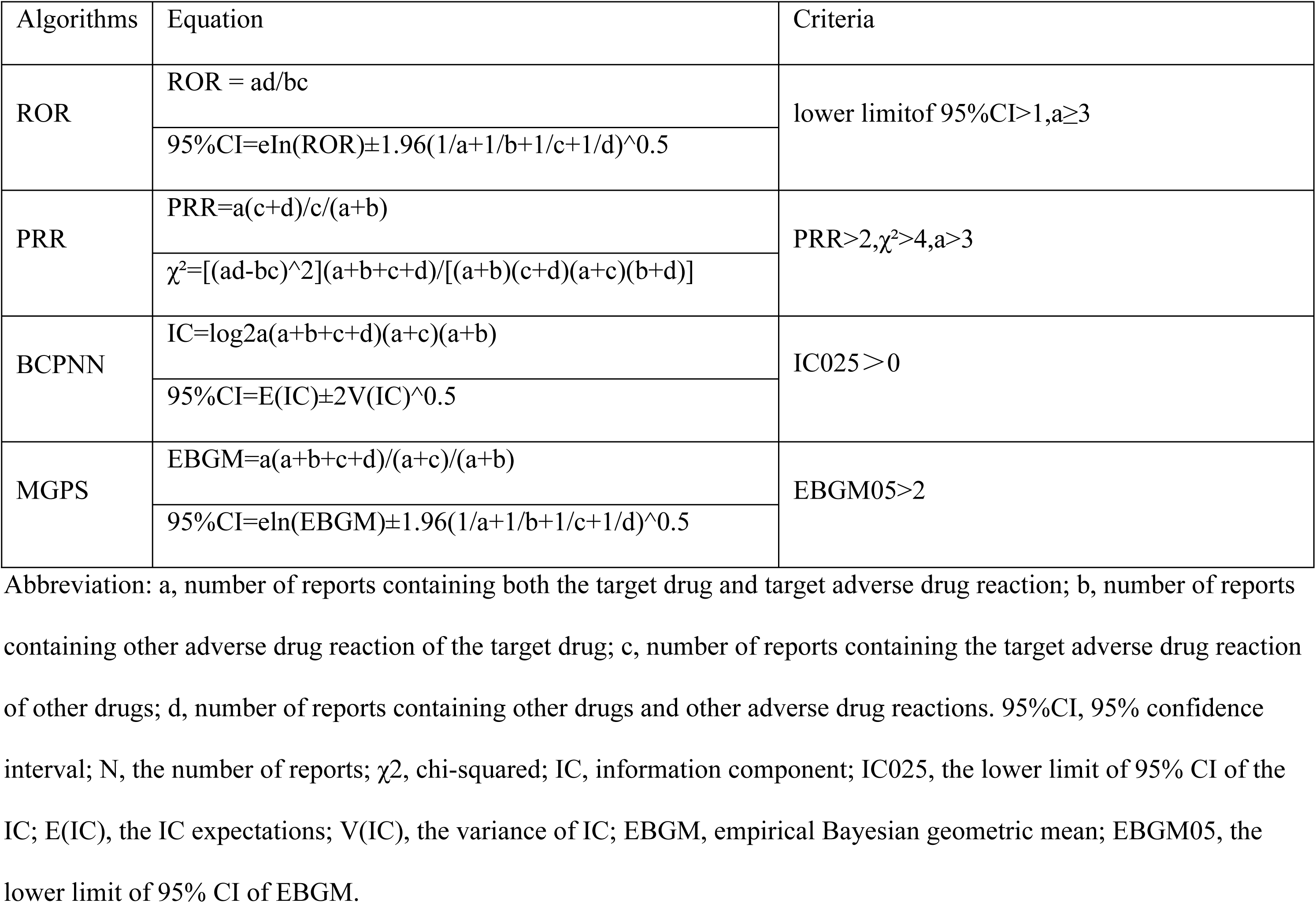
Formulas and signal detection criterias.

Additionally, we performed descriptive statistical analysis to summarize the clinical characteristics of reports with cymbalta-related AEs, including sex, age, weight, reporting year, reporter country, indication, outcome, and time-to-onset (TTO). TTO was defined as the interval from the initiation of cymbalta therapy to the occurrence of an AE; reports with missing or illogical onset data were excluded.

This integrated approach enabled a comprehensive and robust assessment of cymbalta-associated safety signals in a large real-world dataset.

### 2.9 Ethics statement

This study is based on existing publications and public databases; both ethical approval and informed consent have been received by each relevant institutional review committee.

## 3 Results

### 3.1 Drug targets and genetic instruments

After merging and deduplication of targets from traditional Chinese and Western medicines, a total of 572 unique antidepressant drug targets were compiled.

Based on the criteria for pQTLs, 4046 single nucleotide polymorphisms (SNPs) of cis-pQTLassociated to 67 proteins were selected. The F statistic for all instrumental variants exceeds 10, indicating that weak instrument bias is unlikely in our study.

### 3.2 Two-sample MR analysis

Utilizing IVW and Wald ratio methods, along with FDR correction, we identified 15 proteins with significant causal relationships to CAD, as illustrated in Fig. 2.

**Fig. 2.**
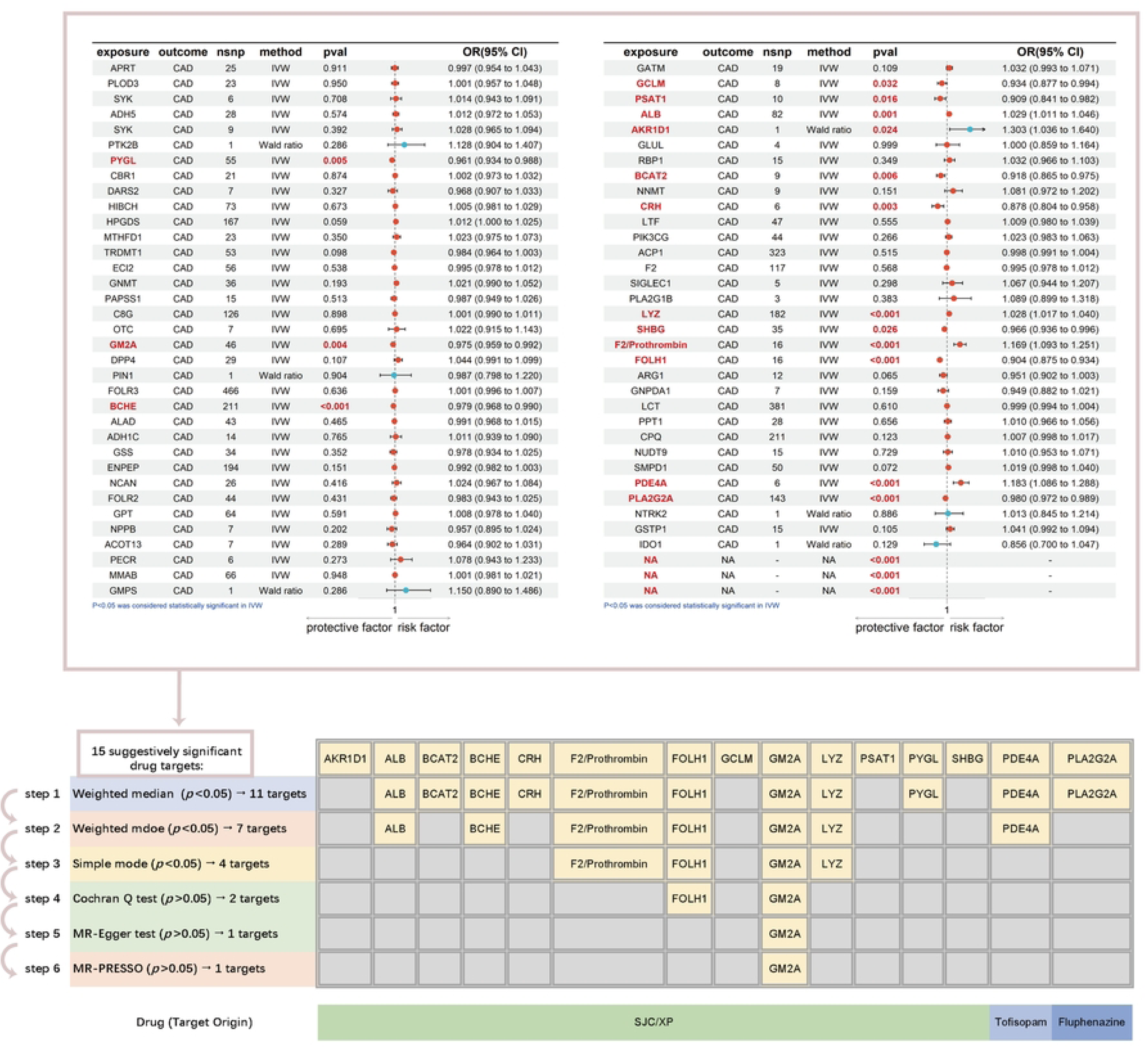
Two-sample MR and sensitivity analyses. All results were derived from the IVW or wald ratio method. The p-values have been adjusted using FDR correction. MR: Mendelian Randomization. IVW: inverse variance weighted FDR: False Discovery Rate. OR: Represents the odds ratio, which quantifies how the odds of the outcome change with an increase in the exposure variable. CAD: Coronary artery disease. IVW: inverse variance weighted. SJC/XP: Shugan Jieyu Capsules or Xiaoyao Pills

Among these, elevated levels of GM2A (OR: 0.975, 95% CI: 0.959–0.992, *P*_FDR_ = 4×10⁻³), PYGL (OR: 0.961, 95% CI: 0.934–0.988, *P*_FDR_ = 5×10⁻³), BCHE (OR: 0.979, 95% CI: 0.968–0.990, *P*_FDR_ < 1×10⁻³), GCLM (OR: 0.961, 95% CI: 0.934–0.988, *P*_FDR_ = 3.2×10⁻²), PSAT1 (OR: 0.909, 95% CI: 0.841–0.982, *P*_FDR_ = 1.6×10⁻²), BCAT2 (OR: 0.981, 95% CI: 0.865–0.975, *P*_FDR_ = 6×10⁻³), CRH (OR: 0.878, 95% CI: 0.804–0.958, *P*_FDR_ = 3×10⁻³), SHBG (OR: 0.966, 95% CI: 0.936–0.996, *P*_FDR_ = 3×10⁻³), FOLH1 (OR: 0.904, 95% CI: 0.875–0.934, *P*_FDR_ < 1×10⁻³), and PLA2G2A (OR: 0.980, 95% CI: 0.972–0.989, *P*_FDR_ < 1×10⁻³) were significantly associated with a decreased risk of CAD. Conversely, higher concentrations of PDE4A (OR: 1.183, 95% CI: 1.086–1.288, *P*_FDR_ < 1×10⁻³), ALB (OR: 1.029, 95% CI: 1.011–1.046, *P*_FDR_ < 1×10⁻³), AKR1D1 (OR: 1.303, 95% CI: 1.036– 1.640, *P*_FDR_ = 2.4×10⁻²), LYZ (OR: 1.028, 95% CI: 1.017–1.040, *P*_FDR_ < 1×10⁻³), and F2/Prothrombin (OR: 1.169, 95% CI: 1.093–1.251, *P*_FDR_ < 1×10⁻³) were linked to an increased CAD risk.

Notably, the majority of significant proteins are drug targets of XJC or XP, except PDE4A and PLA2G2A, which are targeted by tofisopam and fluphenazine, respectively.

The robustness of these findings was supported by sensitivity analyses, including weighted median, weighted mode, and simple mode MR methods (Fig. 2; Fig. 6A), which consistently identified *GM2A* as a robust protective factor. F2/Prothrombin, FOLH1, GM2A also pass these three analyses. However, further quality control—comprising Cochran’s Q test, MR-Egger intercept, and MR-PRESSO—demonstrated that only GM2A met all criteria, showing neither heterogeneity nor horizontal pleiotropy (Fig. 2). Leave-one-out analyses (Fig. 3) confirmed that no single genetic variant unduly influenced causal estimates, and funnel and scatter plots further supported the findings’ reliability (Fig. 4; Fig. 5).

**Fig. 3.**
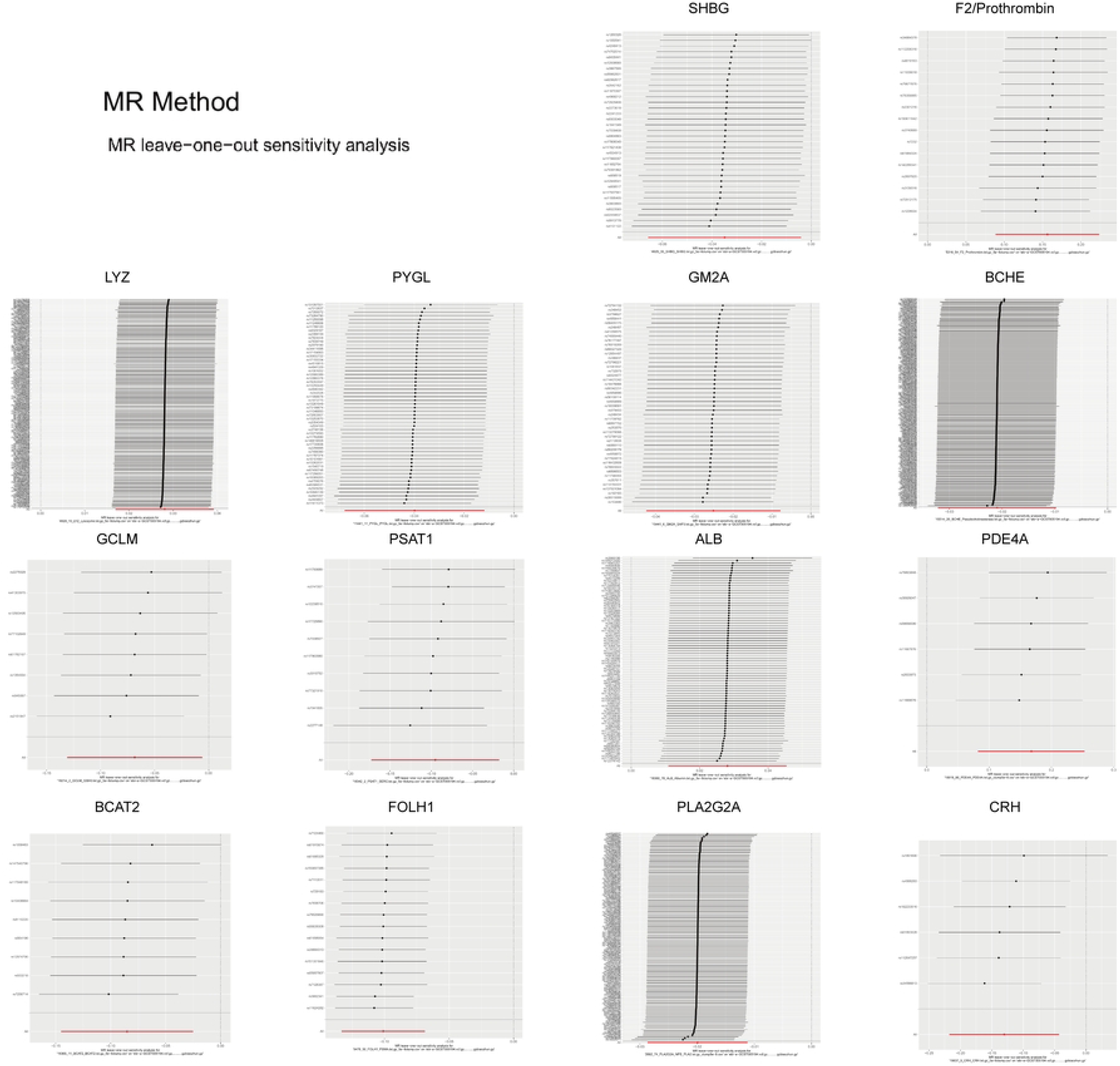
Main leave-one-out sensitivity analysis results

**Fig. 4.**
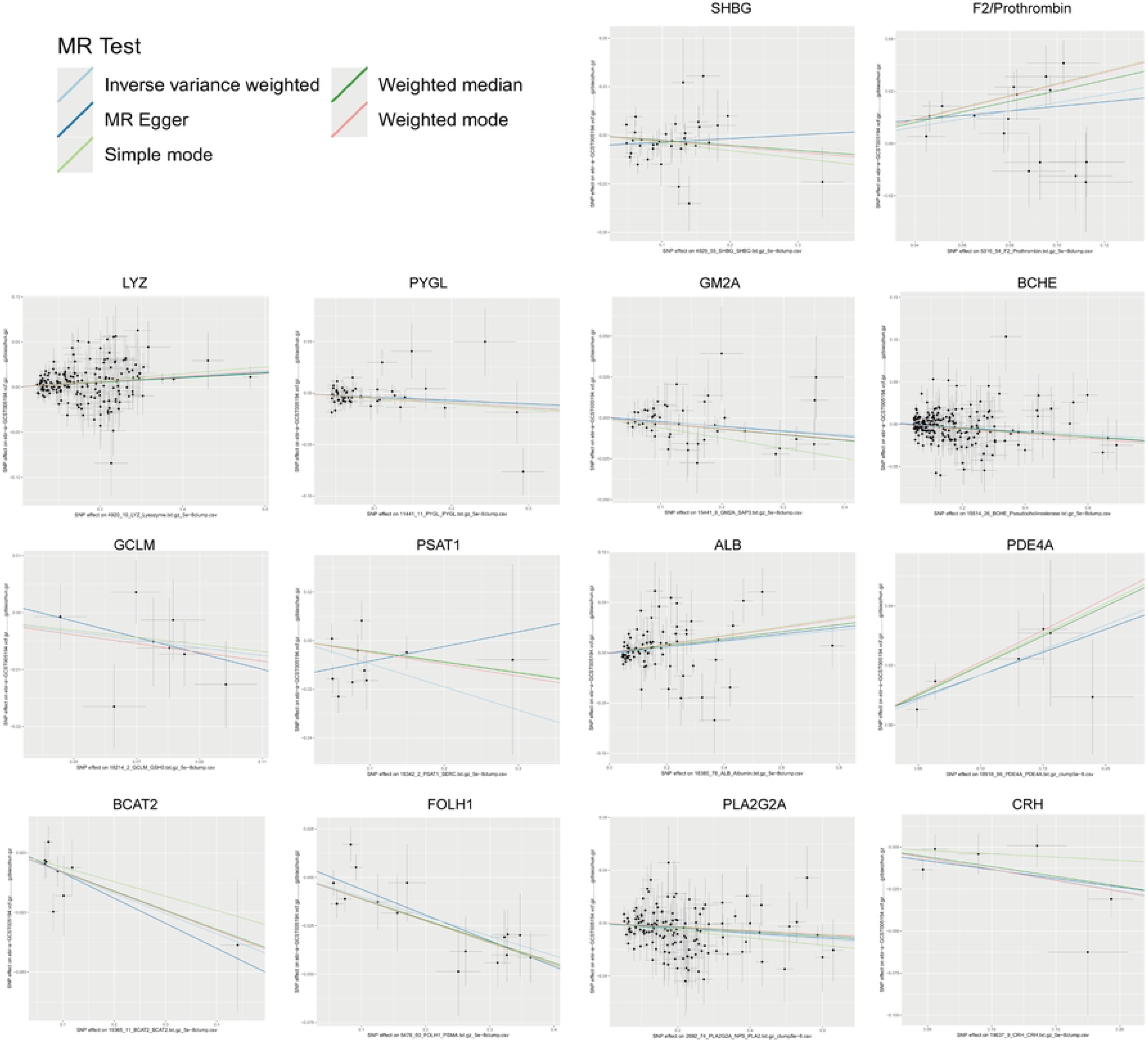
Scatter plot of main Mendelian randomization analyses

**Fig. 5.**
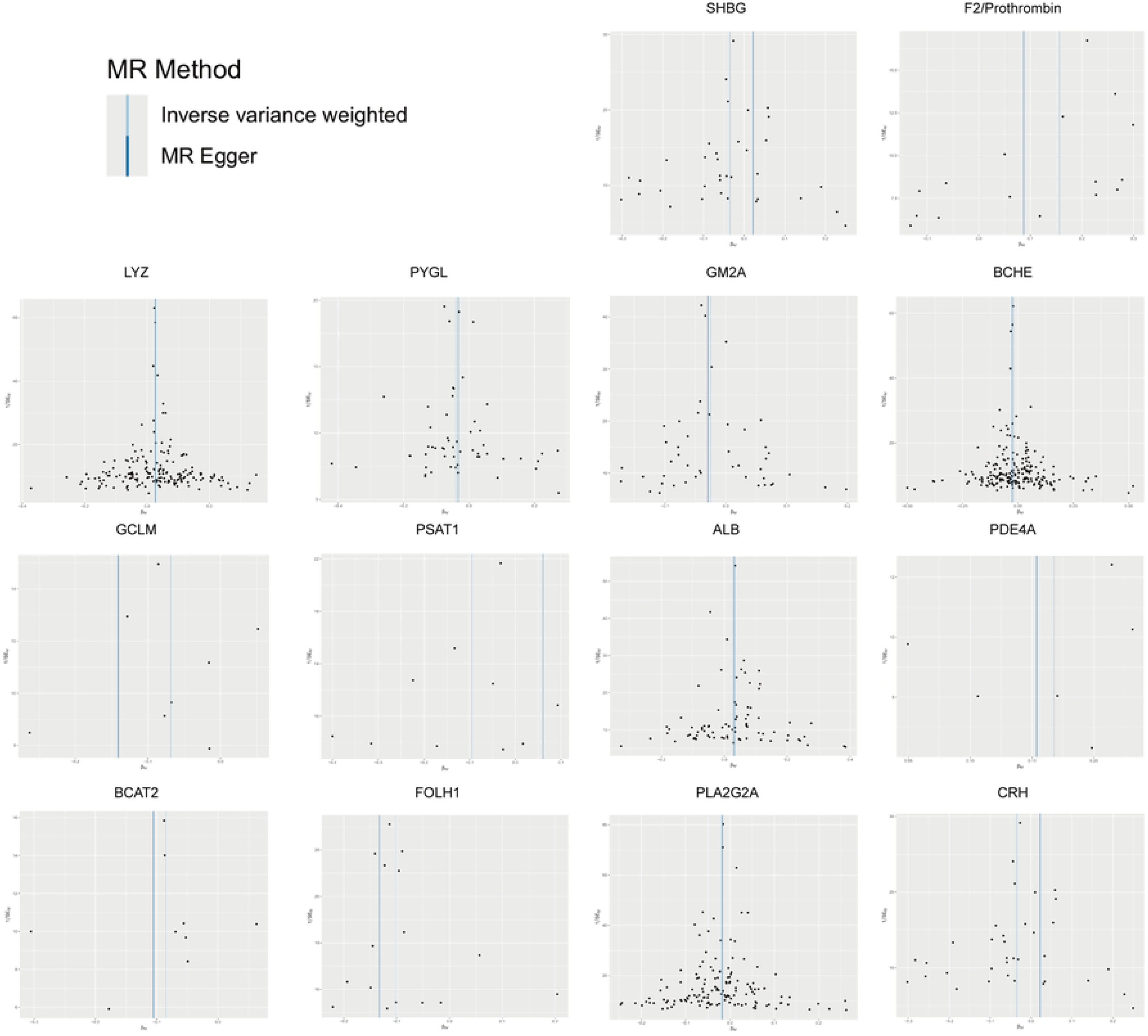
Funnel plot of main Mendelian randomization analyses

**Fig. 6.**
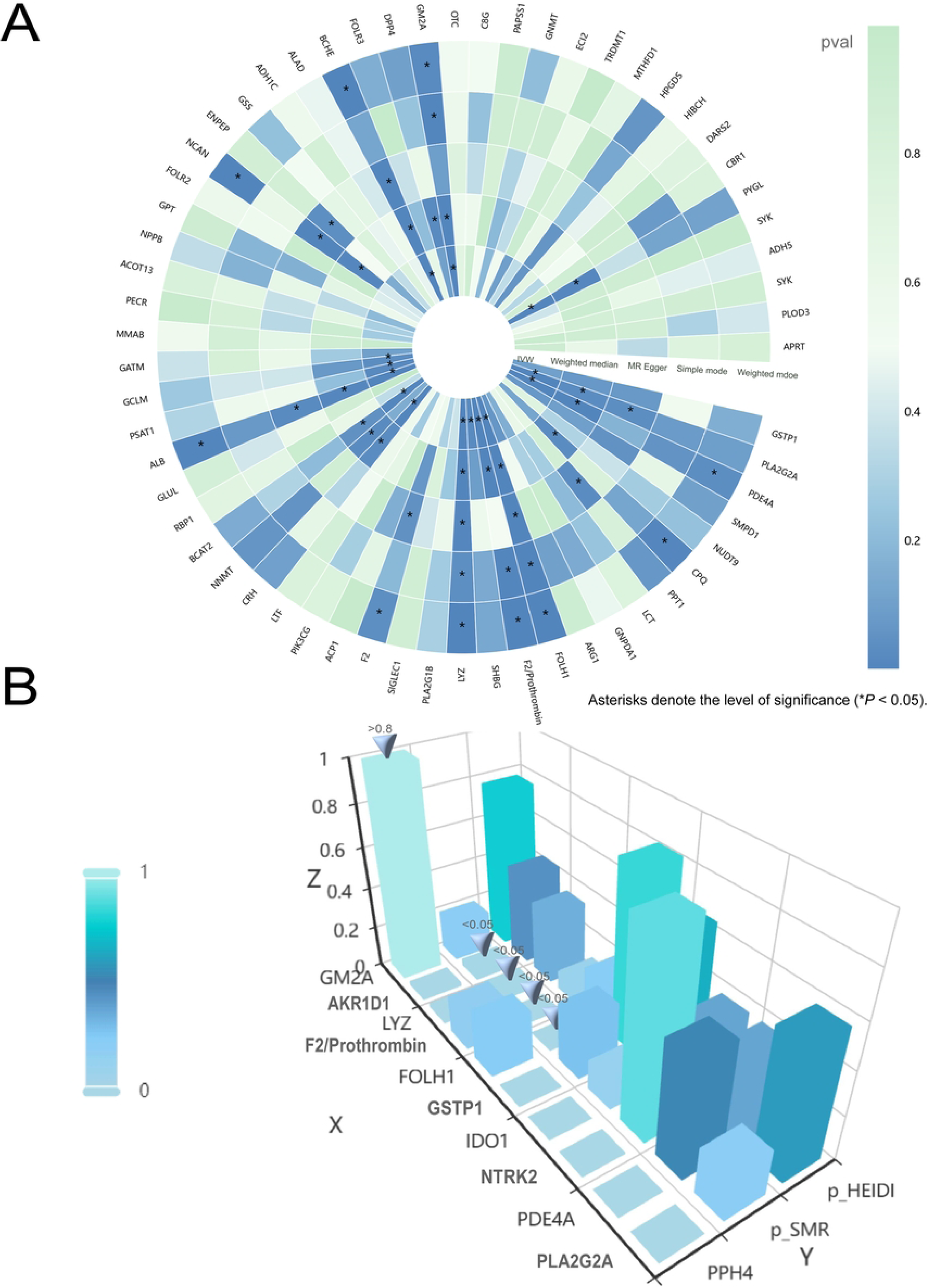
(A) Five methods in two-sample in Mendelian Randomization; (B) Ressults of primary summary dataebased MR (SMR), heterogeneity in dependent instruments (HEIDI) test, and cololization. Asterisks denote the level of significance (\**P* < 0.05).

Collectively, these results highlight GM2A as a key protective protein for CAD and underscore additional candidate proteins as potential therapeutic targets for future investigation.

### 3.3 SMR analysis

In subsequent validation analyses, we performed SMR along with the HEIDI test to validate the results. Of fifteen candidate proteins (AKR1D1, ALB, BCAT2, BCHE, CRH, F2/Prothrombin, FOLH1, GCLM, GM2A, LYZ, PSAT1, PYGL, SHBG, PDE4A, and PLA2G2A), four (F2/Prothrombin, FOLH1, LYZ, and AKR1D1) demonstrated significant associations with CAD (*P*_SMR_ < 0.05 and *P*_HEIDI_ > 0.01) in the deCODE cohort, supporting their potential causal effects (Fig. 6B). Specifically, increased levels of F2/Prothrombin (OR = 1.234, 95% CI: 1.092–1.394; *P*_SMR_ = 0.001; *P*_HEIDI_ = 0.092) and LYZ (OR = 1.026, 95% CI: 1.003–1.050; *P*_SMR_ = 0.027; *P*_HEIDI_ = 0.346) were associated with higher CAD risk, while FOLH1 (OR = 0.893, 95% CI: 0.831–0.959; *P*_SMR_ = 0.002; *P*_HEIDI_ = 0.223) and AKR1D1 (OR = 0.767, 95% CI: 0.598–0.983; *P*_SMR_ = 0.036; *P*_HEIDI_ = 0.441) were protective. Notably, while the direction of effect for F2/Prothrombin, LYZ, and FOLH1 was consistent across _SMR_ and two-sample MR analyses, the direction for AKR1D1 conflicted between methods, warranting further investigation.

Replication analysis in the UKB-PPP dataset reinforced the inverse association of FOLH1 with CAD (OR = 0.829, 95% CI: 0.703–0.978; *P*_SMR_ = 0.026; *P*_HEIDI_ = 0.260), further validating its protective role. BCHE also displayed a protective effect (OR = 0.972, 95% CI: 0.944–1.000; *P*_SMR_ = 0.049; *P*_HEIDI_ = 0.765), which was consistent with two-sample MR, but inconsistent with deCODE-based SMR results.

Unfortunately, no pQTL data for AKR1D1, ALB, BCAT2, F2/Prothrombin, PLA2G2A, PSAT1, PYGL, or SHBG were available in the UKB-PPP dataset, limiting replication analysis for these proteins.

Overall, our multi-cohort SMR investigation highlights FOLH1 as a replicable protective factor for CAD, while other protein associations require further validation in independent datasets.

### 3.4 Bayesian co-localization analysis

To mitigate confounding by LD and bolster causal inference from MR, we performed Bayesian colocalization analysis. Strong evidence indicated a shared causal variant (rs1065853) between GM2A protein levels and coronary artery disease (CAD) risk (PP.H4 = 99.6%). Consequently, GM2A was classified as a tier 1 target. Conversely, AKR1D1 (PP.H4 = 2.0%), LYZ (PP.H4 = 2.1%), F2/Prothrombin (PP.H4 = 18.0%), and FOLH1 (PP.H4 = 23.8%) exhibited limited colocalization evidence with CAD and were designated tier 3 targets. Notably, PLA2G2A and PDE4A associations with CAD may be driven by distinct causal variants (PP.H3 > 0.8). These findings strengthen the evidence for GM2A as a validated therapeutic target for antidepressant intervention in CAD. Results of the colocalization analysis are presented in **Error! Reference source not found.**.

### 3.5 Tissue-specific expression of the key target

Significantly, the association between *GM2A* gene expression and CAD was replicated across six tissues—artery aorta, nerve tibial, thyroid, pituitary, stomach, and pancreas—in the GTEx dataset. SMR analysis revealed that higher *GM2A* expression in all 26 surveyed tissues was inversely associated with CAD risk, but only these six tissues exhibited statistically significant associations (Fig. 7). Specifically, elevated *GM2A* expression was linked to a reduced risk of CAD in the artery aorta (OR = 0.892, 95% CI: 0.813–0.978, *P* < 0.05), nerve tibial (OR = 0.904, 95% CI: 0.825–0.990, *P* < 0.05), thyroid (OR = 0.866, 95% CI: 0.759–0.988, *P* < 0.05), pituitary (OR = 0.927, 95% CI: 0.865–0.994, *P* < 0.05), stomach (OR = 0.915, 95% CI: 0.842–0.994, *P* < 0.05), and pancreas (OR = 0.901, 95% CI: 0.816–0.995, *P* < 0.05), with *P*_HEIDI_ > 0.01 for all.

**Fig. 7.**
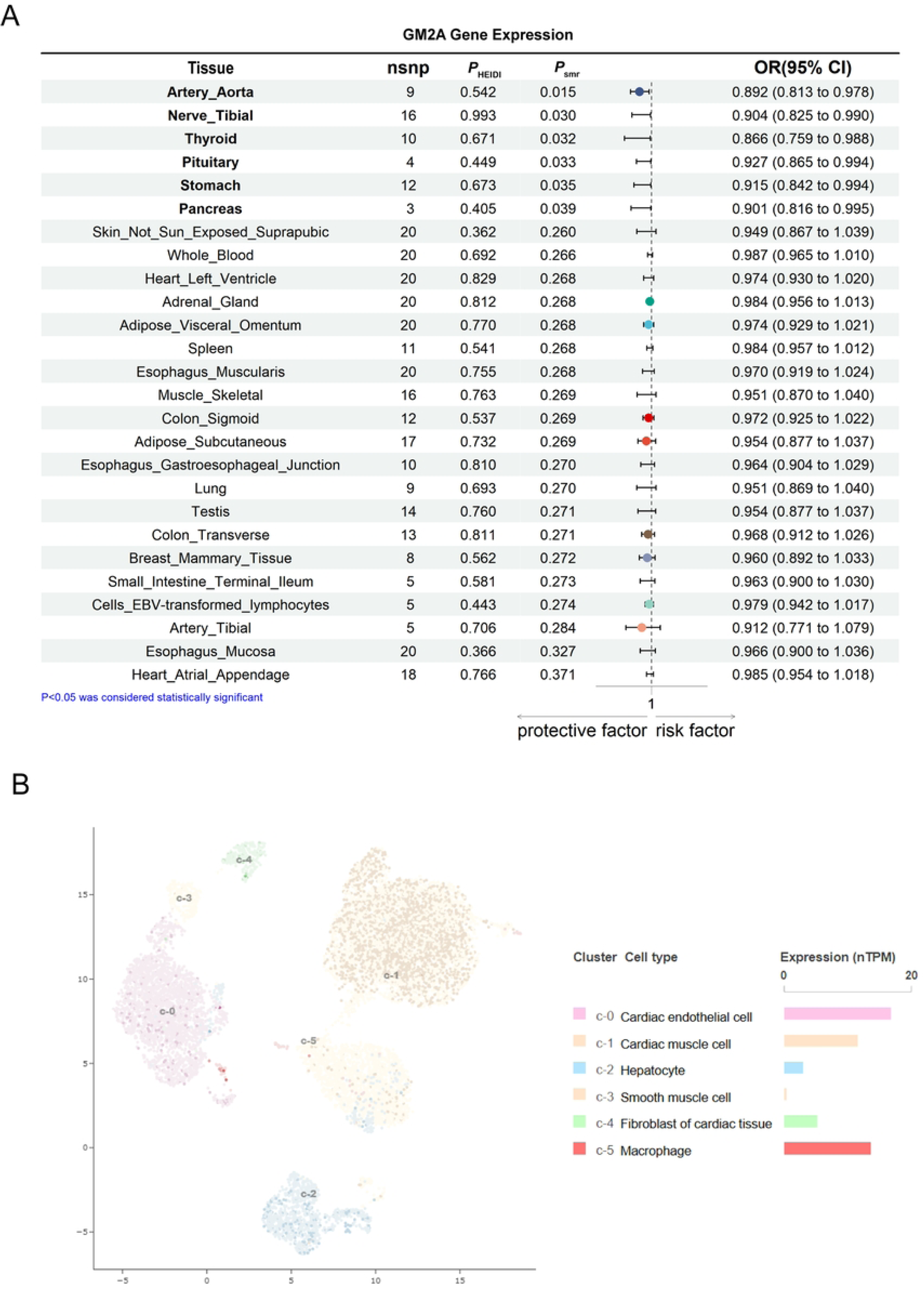
**(A)** Tissue-specific expression of GM2A; (B) Cell type specificity of *GM2A*.

Single-cell RNA sequencing revealed that the *GM2A* gene, which encodes the *GM2A* protein, shows high expression in the caudate, heart muscle, hippocampus, and gallbladder, and moderate expression in the liver, cerebellum, cerebral cortex, stomach, and thyroid gland. At the cellular level, *GM2A* showed specificity for cardiomyocytes, hepatocytes, mitotic cells (heart, thyroid, and stomach), cholangiocytes, visceral adipocytes, lactotropes, somatotropes, glandular epithelial cells, hepatic NK cells, corticotropes, gastric enteroendocrine cells, and chief cells. Among heart tissue cell types (Fig. 7B), *GM2A* transcript levels (nTPM) were highest in cardiac endothelial cells (c-0), minimal in smooth muscle cells (c-3), fibroblasts (c-4), and hepatocytes (c-2), and intermediate in cardiomyocytes (c-1) and macrophages (c-5).

Overall, these findings suggest that tissue- and cell-type-specific expression of *GM2A* may play a protective role in CAD, highlighting its potential as a therapeutic target.

### 3.6 Descriptive analysis

Following the analysis of the causal links between antidepressant targets and CAD, we systematically assessed the potential risks associated with antidepressant treatment in CAD, integrating evidence from pharmacovigilance (FAERS) data to comprehensively evaluate their therapeutic potential. Among available options, cymbalta—a serotonin-norepinephrine reuptake inhibitor (SNRI) considered a favorable second-line antidepressant for cardiac patients—was selected for in-depth analysis due to its dual neurotransmitter mechanism and widespread use in depression and pain, both common in the CAD population .

Finally, a total of 6,391 adverse event (AE) reports associated with cymbalta were retrieved from Q1 2004 to Q3 2024, as shown in Table 2. The temporal distribution of reports demonstrated an initial increase, peaking in 2017 with 2,324 cases, followed by a subsequent decline (**Fig. 8**D). In terms of sex distribution, male patients accounted for the majority of cases (3,828; 59.9%), significantly outnumbering females (1,661; 26.0%), indicating that males may be more susceptible to cymbalta-related AEs.

**Fig. 8.**
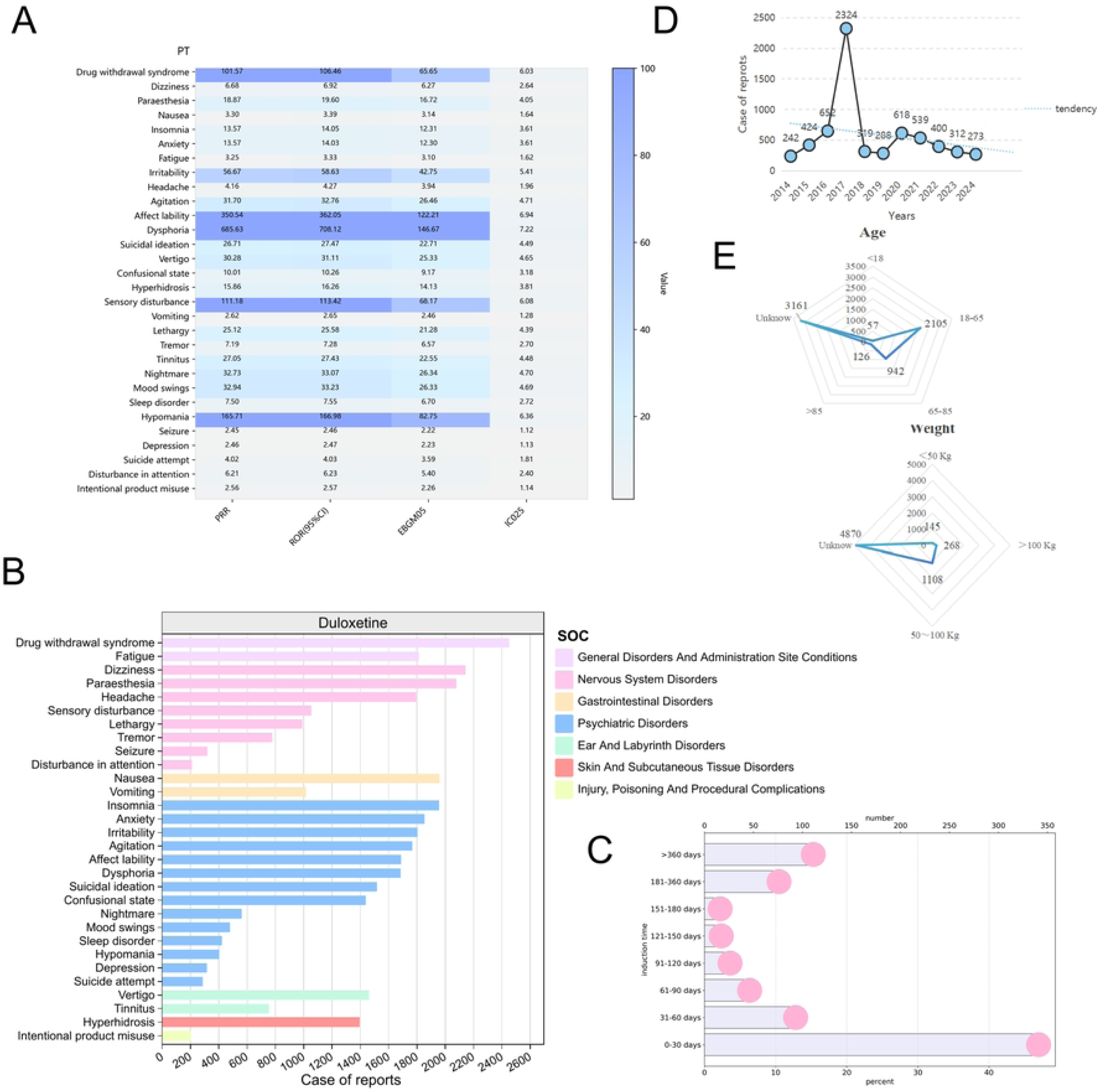
Pharmacovigilance study based on the FAERS database. (A) Top 30 AEs with the highest percentage of signal detection; (B) AEs related to the SOC level. (C) Time-to-onset of AEs. (D) Line chart of reported years. (E) Radar charts of age and weight. FAERS: FDA Adverse Event Reporting System. AEs: adverse events. PTs: preferred terms. ROR: Reporting Odds Ratio. PRR: Proportional Reporting Ratio. EBGM05: Empirical Bayes Geometric Mean, 5th percentile. IC025: Information Component, 2.5% lower confidence bound. SOC: system organ class.

**Table 2.** Clinlcal characteristics of cymbalta adverse event reportsfrom the FAERS database (Q1 2004-Q3 2024)

|  | Cymbalta (n=6391) |  |
| --- | --- | --- |
|  |  | n |
| Sex | Male | 3828 (59.9%) |
|  | Female | 1661 (26.0%) |
|  | Unknow | 902 (14.1%) |
| Age, years | <18 | 57 (0.9%) |
|  | 18-65 | 2105 (32.9%) |
|  | 65-85 | 942 (14.7%) |
|  | >85 | 126 (2.0%) |
|  | Unknow | 3161 (49.5%) |
| Weight | <50 kg | 145 (2.3%) |
|  | >100 kg | 268 (4.2%) |
|  | 50~100 kg | 1108 (17.3%) |
|  | Unknow | 4870 (76.2%) |
| REPORTER COUNTRY | UNITED STATES | 4110 (64.3%) |
|  | UNITED KINGDOM | 401 (6.3%) |
|  | TURKEY | 26 (0.4%) |
|  | THAILAND | 1 (0.0%) |
|  | TAIWAN | 6 (0.1%) |
|  | SWITZERLAND | 36 (0.6%) |
|  | SWEDEN | 33 (0.5%) |
|  | SPAIN | 118 (1.8%) |
|  | SOUTH AFRICA | 7 (0.1%) |
|  | SLOVENIA | 3 (0.0%) |
|  | SAUDI ARABIA | 3 (0.0%) |
|  | ROMANIA | 4 (0.1%) |
|  | PORTUGAL | 19 (0.3%) |
|  | POLAND | 21 (0.3%) |
|  | PHILIPPINES | 1 (0.0%) |
|  | PAKISTAN | 1 (0.0%) |
|  | OMAN | 1 (0.0%) |
|  | NORWAY | 2 (0.0%) |
|  | NETHERLANDS | 21 (0.3%) |
|  | MOROCCO | 1 (0.0%) |
|  | MEXICO | 4 (0.1%) |
|  | KUWAIT | 3 (0.0%) |
|  | KOREA, SOUTH | 7 (0.1%) |
|  | JAPAN | 326 (5.1%) |
|  | ITALY | 109 (1.7%) |
|  | ISRAEL | 5 (0.1%) |
|  | IRELAND | 6 (0.1%) |
|  | INDIA | 11 (0.2%) |
|  | ICELAND | 1 (0.0%) |
|  | HUNGARY | 2 (0.0%) |
|  | GREECE | 17 (0.3%) |
|  | GERMANY | 396 (6.2%) |
|  | FRANCE | 376 (5.9%) |
|  | FINLAND | 4 (0.1%) |
|  | ESTONIA | 1 (0.0%) |
|  | EL SALVADOR | 1 (0.0%) |
|  | DENMARK | 19 (0.3%) |
|  | CROATIA | 11 (0.2%) |
|  | COUNTRY NOT SPECIFIED | 22 (0.3%) |
|  | COSTA RICA | 1 (0.0%) |
|  | CHINA | 9 (0.1%) |
|  | CANADA | 175 (2.7%) |
|  | BULGARIA | 1 (0.0%) |
|  | BRAZIL | 6 (0.1%) |
|  | BELGIUM | 9 (0.1%) |
|  | BARBADOS | 2 (0.0%) |
|  | AUSTRIA | 17 (0.3%) |
|  | AUSTRALIA | 35 (0.5%) |
| Indication | DEPRESSION | 1217 (19.0%) |
|  | ANXIETY | 159 (2.5%) |
|  | BACK PAIN | 109 (1.7%) |
|  | FIBROMYALGIA | 227 (3.6%) |
|  | PAIN | 577 (9.0%) |
|  | NEURALGIA | 175 (2.7%) |
|  | others | 3927(61.5%) |

Demographic analysis indicated that adults aged 18–65 years constituted the largest affected group (2,105; 32.9%), as illustrated in **Fig. 8**E, while individuals aged 65–85 years accounted for 14.7% (942 cases). After excluding cases with unknown information, it was found that the majority of affected individuals had a body weight between 50 and 100 kg (**Fig. 8**E). Geographically, the United States contributed the largest proportion of AE reports (4,110; 64.3%), followed by the United Kingdom (401; 6.3%).

Analysis of indications revealed that cymbalta was most frequently prescribed for depression (1,217; 19.0%), followed by pain (577; 9.0%), with other common indications including anxiety, back pain, fibromyalgia, and neuralgia. This distribution underscores cymbalta’s primary use in managing depressive disorders and various pain conditions.

Notably, the median time-to-onset (TTO) for cymbalta-related AEs was 255 days, with some cases exceeding 360 days (**Fig. 8**C). This indicates a substantial delay between drug initiation and AE reporting, potentially hindering the prompt identification and management of safety signals.

In summary, cymbalta-related AE reports in the FAERS database predominantly involved male patients and adults aged 18–65 years, with the majority reported from the United States. The observed time lag in AE onset underscores the need for improved vigilance and timely reporting to facilitate early detection of potential safety concerns.

These findings highlight the necessity for continued pharmacovigilance, particularly for high-risk populations, to optimize the safe use of cymbalta.

### 3.7 Disproportionality analysis

A total of 1,961 preferred terms (PTs) were identified as AEs related to cymbalta. Among these, the top 30 most frequently reported AEs, visualized as a heatmap in **Fig. 8**A, included both expected events (e.g., somnolence, seizure, vomiting) and unexpected events not commonly noted in the label or clinical trials (e.g., drug withdrawal syndrome, dizziness, paraesthesia, insomnia, anxiety, fatigue, irritability, headache, agitation). Higher ROR and PRR values indicate stronger AE signals, reflecting a greater statistical association between the drug and the specific adverse event. The most significant safety signals were detected for dysphoria (n = 1,684; ROR 708.12, 95% CI 101.35–111.82; IC 7.32), affect lability (ROR 362.05, 95% CI 334.61–391.75; IC 7.03), hypomania (n = 403; ROR 166.98, 95% CI 146.42–190.42; IC 6.53), sensory disturbance (n = 1,053; ROR 113.42, 95% CI 105.18–122.3; IC 6.18), and drug withdrawal syndrome (n = 2,452; ROR 106.46, 95% CI 101.35–111.82; IC 6.1). Seizure, although expected, demonstrated the lowest signal (n = 319; ROR 2.46, 95% CI 2.2– 2.75; IC 1.26).

At the system organ class (SOC) level (**Fig. 8**B), Psychiatric Disorders accounted for the largest number of reports (n = 18,447; ROR 11.86, 95% CI 11.65–12.08; IC 2.97), while Ear and Labyrinth Disorders demonstrated the strongest signal intensity (n = 2,274; ROR 13.27, 95% CI 12.71–13.86; IC 3.59). Notably, robust associations were also observed for Nervous System Disorders (n = 16,000; ROR 3.35, 95% CI 3.28–3.42; IC 1.49) and Gastrointestinal Disorders (n = 4,978; ROR 1.15, 95% CI 1.11–1.18). Some SOCs lacked statistical significance as their ROR 95% CIs included or fell below 1.

## 4 Discussion

CAD remains the leading global cause of mortality, with depression recognized as an independent risk factor that exacerbates it by promoting sympathetic overactivation and increasing cardiac burden (3). Depressive symptoms affect a significant portion of patients hospitalized for coronary events, complicating management due to the adverse cardiovascular effects and dependencies linked to conventional antidepressants (15). TCM, exemplified by Ginkgo biloba dropping pills (GBDP), approved for concurrent cardiovascular and neuropsychiatric symptom management, offers a complementary approach aligned with psycho-cardiology principles (16). Therefore, antidepressant drug targets were comprehensively collected from both TCM and Western antidepressants in this study. We used MR to investigate the potential of traditional Chinese and Western antidepressants for treating CAD and analyzed the safety of representative antidepressants using the FAERS database.

In this MR study, we aimed to elucidate the causal role of antidepressant drug targets in CAD risk. Our findings reveal significant associations between specific antidepressant targets and CAD outcomes, suggesting that neuropsychiatric factors contribute to CAD pathogenesis and that antidepressant medications might possess therapeutic potential as anti-CAD agents. Unlike prior studies that primarily examined the influence of antidepressant use on CAD progression, our analysis provides novel insights into how distinct antidepressant targets may differentially impact CAD onset. Using a two-sample MR approach, we identified several drug targets with potential causal effects on CAD risk. Targets associated with the SJC/XP, including GM2A, PYGL, BCHE, BCAT2, SHBG, CRH, FOLH1, GCLM, and PSAT1, as well as the fluphenazine target PLA2G2A, were linked to reduced CAD risk. Conversely, targets of tofisopam (PDE4A) and additional SJC/XP targets (AKR1D1, ALB, LYZ, and F2/Prothrombin) were associated with increased CAD risk. Notably, fluphenazine and tofisopam act as inhibitors on PLA2G2A and PDE4A, respectively; thus, fluphenazine’s inhibitory action may elevate CAD risk by suppressing protective PLA2G2A expression, whereas tofisopam’s inhibition of PDE4A may reduce CAD risk. The precise effects of traditional Chinese medicine on targets like GM2A and others remain unclear but may involve dual-directional regulatory mechanisms that restore physiological balance in various systems.

The strength of our instrumental variables was supported by F statistics exceeding 10, mitigating concerns of weak instrument bias. While all 15 identified protein targets demonstrated significance after false discovery rate correction using IVW MR, the potential for instability warranted rigorous sensitivity analyses. To address this issue, we employed multiple complementary methods to validate causal inference robustness: The weighted median estimator provides a consistent causal effect estimate even if up to 50% of instruments are invalid by focusing on the median of weighted effects, thus reducing outlier influence (17). Similarly, the weighted mode estimator identifies the most frequent effect weighted by instrument precision, offering robustness when most instruments share a common effect (17). In contrast, the simple mode estimator detects predominant effects without weighting, useful for finding consensus amid heterogeneity (17). To evaluate variability across instruments, Cochran’s Q test detects heterogeneity likely from pleiotropy or invalid instruments (18). MR-Egger regression further tests for directional pleiotropy via its intercept, where significance indicates pleiotropic bias (18). Lastly, MR-PRESSO identifies and corrects pleiotropy-induced outliers, enhancing estimate stability (18).

Among the 15 targets, GM2A emerged as the most robustly supported by these sensitivity analyses, and the potential value of the other 14 targets warrants further investigation.

To further disentangle pleiotropy from linkage disequilibrium confounding, we applied SMR alongside HEIDI testing. A HEIDI *p*-value > 0·01 indicates a low likelihood of linkage-induced bias (19). Colocalization analysis complemented these methods, with PP.H4 above 0·8 was considered strong evidence for colocalization (20). A PP.H4 greater than 0.8, as seen with GM2A, reinforces confidence in shared causality with CAD, designating GM2A as a tier 1 target under our criteria.

Multi-cohort SMR analysis identified FOLH1 as a replicable protective factor for CAD, consistent with our two-sample MR results. FOLH1 encodes a folate-hydrolyzing transmembrane glycoprotein implicated in regulating homocysteine and lipid metabolism, known contributors to CAD (21). Although polymorphisms in FOLH1 correlate with higher homocysteine levels and dyslipidemia—risk factors that promote endothelial dysfunction and thrombosis—our MR findings suggest increased FOLH1 protein expression paradoxically reduces CAD risk. This discrepancy challenges prior observational evidence and pharmacological assumptions, necessitating further mechanistic studies to clarify FOLH1’s role in CAD pathogenesis.

GM2A, encoding the GM2 activator protein, is a lysosomal glycosphingolipid transporter critical for ganglioside degradation and lipid trafficking (22). It is predominantly expressed in macrophages and various cardiovascular cell types (23). Our SMR analyses consistently associated elevated GM2A expression in the aorta with reduced CAD risk, underscoring its protective function. Single-cell RNA sequencing revealed GM2A expression specificity in cardiomyocytes, cardiac endothelial cells, and macrophages, highlighting its relevance in cardiovascular lipid homeostasis.

Oxidized low-density lipoprotein (oxLDL) induces GM2A upregulation in macrophages, a response traditionally thought to exacerbate glycosphingolipid accumulation and promote CAD (22). Our data challenge this paradigm, proposing that increased GM2A expression facilitates adaptive glycosphingolipid transport and clearance, thereby mitigating intracellular lipid toxicity and atherogenesis. This novel interpretation positions GM2A as a promising therapeutic modulator of lipid metabolism to reduce CAD risk.

Furthermore, our tissue-specific analyses revealed that elevated GM2A expression in the thyroid, pituitary, stomach, and pancreas is associated with a reduced risk of CAD. This association may reflect the involvement of these tissues in key mechanisms influencing CAD pathophysiology. For instance, thyroid hormones critically regulate cardiac function and vascular tone, while pituitary function modulates the hypothalamic-pituitary-adrenal (HPA) axis, which affects systemic inflammation and stress responses implicated in CAD (24). The stomach and pancreas contribute to metabolic regulation and inflammatory status, both relevant to atherosclerosis development (25). Despite these plausible links, the precise biological pathways through which GM2A expression in these tissues confers cardioprotective effects remain unexplored. Further mechanistic studies are warranted to elucidate the role of GM2A in modulating CAD risk within these specific tissue contexts.

Additionally, this study also leveraged the FAERS database to analyze AEs related to cymbalta since its FDA approval in 2004. Our findings corroborate known AEs documented in prescribing information, including somnolence, seizure, and vomiting, while revealing additional safety signals across diverse categories such as general disorders, nervous system, gastrointestinal, psychiatric, ear and labyrinth, dermatologic, and procedural complications.

Notably, despite a higher prevalence of depression in females, fewer female AE reports emerged, likely reflecting their generally better antidepressant response. The 18–65 age group exhibited the highest proportion of AE reports. Reports from the United States predominated, consistent with FAERS being a US-centric database.

Our data confirmed cymbalta’s primary indications for depression, anxiety, and various pain conditions, aligning with its approved labeling. A pronounced drug withdrawal syndrome was observed, consistent with prior systematic reviews indicating that SNRI discontinuation commonly triggers withdrawal symptoms (26). Among AEs, dizziness was prominent, possibly attributable to cymbalta’s modulation of serotonin and norepinephrine reuptake elevating central neurotransmitter levels, impacting autonomic function, and causing mild tachycardia and orthostatic hypotension (27).

Nervous system disorders were frequently reported, with paresthesia commonly observed during treatment or tapering phases, likely reflecting altered neurotransmitter-mediated sensory processing and nervous system adaptation. Gastrointestinal AEs, particularly nausea, may result from increased serotonin stimulating 5-HT3 receptors, provoking emetic responses typically resolving over time (27). Additional AEs such as insomnia, anxiety, fatigue, irritability, headache, and agitation were detected with notable frequency in FAERS, despite being underrepresented in clinical trials, emphasizing the complexity of nervous system-related adverse effects. Time-course analysis revealed that most AEs clustered within the first month of treatment, underscoring the vital importance of early vigilant monitoring to promptly identify and manage emerging symptoms.

However, the study has several limitations. First, the drug target MR simulates lifetime low-dose drug exposure under idealized conditions. The implications of short-term drug administration on CAD remain unclear, which may not reflect the complexities of real-world environments; thus, clinical trials remain essential for validating drug efficacy. Second, MR assesses the impact of individual druggable gene expression in isolation, while many drugs function through the interplay of multiple targets. Third, data from the Decode and UKB-PPP cohorts, predominantly consisting of European populations, may introduce bias due to population heterogeneity, necessitating the inclusion of diverse genetic backgrounds in future GWAS. Furthermore, RCTs provide stronger causal inferences than MR, indicating the need for high-quality clinical trials to support findings related to GM2A and CAD causation.

Despite these limitations, the study also presents notable advantages. It establishes a cost-effective and efficient method for identifying drug targets, unlike traditional approaches. MR and colocalization analyses effectively assess potential causal influences of druggable targets on ALD, utilizing genetic variation as proxies for exposure, thereby reducing confounding factors inherent in other study designs. Notably, using pQTLs strengthens the causal link between genetic variation and protein levels, enhancing drug target validation. The study maintained methodological consistency by focusing on a homogeneous ancestry group and employed various sensitivity analyses to mitigate bias and confounding influences, ensuring robust causal relations. Despite challenges, pharmacovigilance remains vital for monitoring medication safety.

In conclusion, our study provides compelling evidence that specific antidepressant targets, notably GM2A, may modulate CAD risk through complex mechanisms involving lipid metabolism and neuropsychiatric pathways. Robust sensitivity analyses and colocalization support GM2A as a promising therapeutic target, opening avenues for repurposing antidepressants and traditional medicines in CAD prevention and treatment. Nonetheless, significant safety concerns associated with agents such as cymbalta underscore the need for vigilant clinical monitoring and careful management to optimize patient outcomes. These findings show both the possible benefits and safety issues of using antidepressants for CAD, highlighting the need for more research and clinical testing.

## Data Availability

The data underlying the results presented in the study are available from https://gwas.mrcieu.ac.uk/, https://registry.opendata.aws/ukbppp/, https://epigraphdb.org/, http://www.tcmip.cn/ETCM/,https://go.drugbank.com/unearth/q?searcher=indications&query=depression&button=

## Acknowledgments

This research utilized publicly accessible genome-wide summary statistics from the EpiGraphDB, deCODE, and the UKB-PPP study. We express our gratitude to all the consortia for granting public access to the summary statistics and also acknowledge the contributions of the participants in these studies.

## Author contributions

Conceptualization: Yaojian Wang, Zhidie Jin

Data curation: Yaojian Wang, Zhidie Jin

Formal analysis: Yaojian Wang, Zhidie Jin

Funding acquisition: Yaojian Wang, Zhidie Jin

Investigation: Yaojian Wang, Zhidie Jin

Methodology: Yaojian Wang, Zhidie Jin

Project administration: Yaojian Wang, Zhidie Jin

Resources: Yaojian Wang, Zhidie Jin

Software: Yaojian Wang, Zhidie Jin

Supervision: Yuerong Jiang, Keji Chen

Validation: Yuerong Jiang, Keji Chen

Visualization: Yaojian Wang, Zhidie Jin

Writing – original draft: Yaojian Wang, Zhidie Jin

Writing – review & editing: Yaojian Wang, Zhidie Jin

